# Impacts of warning labels on ultra-processed foods among Latino adults: A randomized trial

**DOI:** 10.64898/2026.03.18.26348497

**Authors:** Lindsey Smith Taillie, Violet Noe, Mrignyani Sehgal, Aline D’Angelo Campos, Anna Grummon, Jennifer Falbe, Aviva A. Musicus, Carmen E. Prestemon, Cristina J. Y. Lee, Marissa G. Hall

**Author notes:** Corresponding author: Violet Noe, MPH, RDN, Project Manager, Global Food Research Program, The University of North Carolina at Chapel Hill, 123 West Franklin St, Chapel Hill, NC 27516.

## Abstract

**Introduction:** Ultra-processed foods (UPFs), defined as foods in group 4 of the NOVA classification system, are a key contributor to chronic disease in the United States. Front-of-package warning labels (“warnings”) offer a promising strategy to help Americans reduce consumption of UPFs. Requiring warning labels on UPFs could help reduce consumption of these foods. However, the effects of UPF warnings are largely unknown. The impact of warning labels on UPFs among Latino adults was examined.

**Study design:** Online randomized trial.

**Setting/participants:** 4,107 Latino adults (49% limited English proficiency) in the US.

**Intervention:** Participants viewed one of three labels: *control labels* displaying barcodes; *identity warnings* stating “WARNING: Ultra-processed food”; or *health warnings* stating “WARNING: Consuming ultra-processed food and drinks can cause weight gain, which increases the risk of obesity and type 2 diabetes”.

**Main outcome measures:** Participants viewed four UPF products displaying their randomly assigned labels. Participants indicated whether the product was UPF (primary outcome) and rated perceived healthfulness of the product, intentions to purchase the product, and perceived message effectiveness (secondary outcomes).

**Results:** Identity warnings (70% correct) and health warnings (67% correct) both led to higher correct identification of UPF compared to control labels (54%, *p<.001),* with the identity warning having a larger impact than the health warning (*p*=.007). Compared to the control label, the identity warning and health warning both elicited higher perceived message effectiveness and lower perceptions of healthfulness and purchase intentions (*p*<.001 for all outcomes) with no significant differences between UPF labels. The impact of the health warning label (vs. the control label) on correct identification of UPF was greater for participants with high education (*p*=0.012) compared to those with low education, and participants with limited English proficiency (*p*=0.001).

**Conclusions:** UPF warnings may help consumers identify UPFs and influence product perceptions and intentions.

**Trial registration:** NCT06296355.

## Introduction

Consumption of ultra-processed foods (UPFs), defined as foods in category 4 on the NOVA classification system, is highly prevalent in the United States. UPFs are manufactured to be convenient, durable, tasty, and profitable by combining substances extracted from foods with additives such as flavors, colors, or emulsifiers.^1^ Examples of UPFs include cookies, corn chips, ready-to-eat meals, candy, and soft drinks. These foods make up 50% of food purchases and contribute to more than half of the estimated daily calories consumed in the US.^1,2^ Consumption of UPFs, which are often high in calories, added sugars, sodium, and saturated fats, has been associated with numerous health risks including overweight and/or obesity,^3–5^ diabetes,^4,6^ hypertension and cardiovascular diseases,^4,5,7^ mental health disorders,^8^ and all-cause mortality.^5,9^ Randomized trials also demonstrate that diets high in UPFs result in higher energy intake and weight gain compared to diets primarily consisting of unprocessed whole foods.^10,11^ The pervasiveness of UPF consumption and the consistent link between UPF consumption and negative health outcomes point to a need for public policies to help consumers identify and reduce consumption of UPFs.

Front-of-package labels are a promising strategy to help consumers more easily identify UPFs—a key step on the pathway to reducing consumption.^12^ A robust body of research shows that clear, easy-to-understand front-of-package warning labels (“warnings”) about a food’s nutrient content or health effects helps consumers identify unhealthy products,^12–15^ influences perceptions of healthfulness and intentions to purchase these products,^12,16–21^ and reduces purchases.^19,20,22^ Improving consumers’ ability to identify products may be especially important for UPFs, given that ultra-processing is a complex construct involving ingredients and industrial processes—information that is either not conveyed on the package or is difficult to comprehend. Research shows that many consumers lack awareness of the extent of food processing and its health implications, limiting their ability to make informed dietary choices.^23–28^ Yet, there is limited data about whether UPF warnings effectively improve consumers’ ability to identify UPFs. Additionally, UPF warnings are timely and relevant as interest in regulating UPF products grows among American policymakers. In 2025 alone, two dozen state bills targeting UPFs have been introduced, including at least 6 bills requiring warning labels.^29^ At the federal level, a bill to implement required warning labels on UPFs was introduced in the Senate in 2024.^30^

Also unknown is the warning design that is likely to be most effective at helping consumers identify UPFs. Prior research on foods high in nutrients of concern indicates that warnings that communicate about health effects (“health warnings”) tend to have a larger impact than warnings that simply identify products as high in given nutrients of concern (“identity warnings”).^19,31,32^ However, it is unclear whether this trend also applies to UPFs.^25^ In addition, it is unclear whether UPF warnings may be less effective for populations that may face higher barriers to understanding text-based labels, including those with lower education or those with limited English proficiency.

The objective of this study was to examine the impact of a health warning and an identity warning on consumers’ ability to identify UPFs. The study focused specifically on Latino adults because this demographic makes up a high proportion of the U.S. population, at 19%,^33^ and experiences more diet-related conditions than non-Hispanic white and higher-English proficiency populations,^34–39^. This also allowed us to explore whether the effects of UPF labels differed by education as well as language proficiency.

## Methods

### Participants

This study was conducted as an ancillary study of a larger study focused on nutrient labels for added sugar, sodium, and saturated fat.^40^ In August and September of 2024, the panel company ThinkNow recruited a national convenience sample of 4,107 U.S. Latino adults of parental age (18-55 years). Participants were eligible if they were aged 18 years or older, identified as Hispanic or Latino, and currently resided in the U.S. The panel company employed quotas to recruit a sample in which approximately 50% of individuals had limited English proficiency, defined as reporting speaking English “not at all” or “not well” (vs. “well” or “very well”) using a measure adapted from the U.S. Census American Community Survey.^41^ This study was reviewed and approved by the Institutional Review Board at the University of North Carolina, Chapel Hill (#24-0300). All participants provided online written informed consent.

### Stimuli

#### Products

We selected four products from frequently consumed categories (yogurt, fruit drink, breakfast cereal, salty snacks) (**Supplementary Figure 1**). All products selected were considered ultra-processed according to the NOVA classification system.^1^ Products were selected from brands that the study team considered to be well-known and popular among U.S. consumers. The selected products were also ones that consumers may view as relatively healthy and therefore may not readily identify as UPFs. All selected products contained pre-existing messages on the packaging that communicated an element of healthfulness – for example, “fat free” or “zero sugar”.

#### Labels

The study tested one identity warning (“WARNING: Ultra-processed food”) and one health warning (“WARNING: Consuming ultra-processed food and drinks can cause weight gain, which increases the risk of obesity and type 2 diabetes”) (**Supplementary Figure 2**). The text and visual design of the warnings were based on previous research on front-of-package warning labels.^31,42,43^ In previous studies,^20,44^ a barcode control label was used to control for the effects of placing an additional label on the package, including any branding obscured by the labels.

### Procedures

Participants completed an online survey using the Qualtrics survey platform. Participants had the option to complete the survey in either English or Spanish; however, all labels and stimuli were in English (since this is how products and labels would look in the US market). After screening for eligibility and providing informed consent, participants completed the main experimental task for the parent study^40^ before being randomized for this experiment. A parallel trial design was used with participants randomized to see one of three labels on product images: the identity warning label, the health warning label, or the barcode control label. The Qualtrics randomizer function was used to randomize participants to one of these three conditions in a 1:1:1 ratio. Participants then viewed the products displaying their randomly assigned label and answered questions about each product, one at a time. The products were shown in random order. At the end of the survey, participants viewed their randomly assigned label by itself (not applied to a product) and answered an additional question about the label.

### Measures

#### Primary outcome

The primary outcome was the correct identification of the product as ultra-processed. This was selected as the primary outcome given the primary goal of front-of-package warning labels is to help consumers identify unhealthy products, and that this is also a key step on the pathway from label exposure to behavioral change.^12^ After viewing each product, participants were asked, “Do you think this product is ultra-processed?”. Response options included “No”, “Yes”, and “I’m not sure”. For analysis, the response options “No” and “I’m not sure” were combined, since only “Yes” reflects correct identification.

#### Secondary outcomes

The secondary outcomes included the perceived healthfulness of the product, intentions to purchase the product, and perceived message effectiveness (PME) to discourage consumption of UPFs. Questions to measure secondary outcomes were adapted from previous studies.^43,45,46^ Perceived healthfulness was measured with the question, “How good or bad for your health would it be to consume this product every day?”.^45^ Response options ranged from “Very bad” (coded as 1) to “Very good” (coded as 5). Purchase intentions were measured with the question, “How likely would you be to purchase this product in the next week, if it were available?”.^43^ Response options ranged from “Not at all likely” (coded as 1) to “Extremely likely” (coded as 5). Finally, PME was measured using the question “How much does this message discourage you from wanting to consume an ultra-processed food or drink?” adapted from the UNC PME scale,^46^ with response options ranging from “Not at all” (coded as 1) to “Very much” (coded as 5).

### Statistical analysis

The study’s design, hypotheses, and analytic plan were pre-registered on ClinicalTrials.gov (#<u>NCT06296355</u>) on February 28, 2024 prior to beginning data collection. The prediction was that the correct identification of the products as UPF would be highest for participants assigned to the identity warning label, followed by the health warning label, and then the control label. For the secondary outcomes, we predicted that perceptions of healthfulness and intentions to purchase the products would be lower and perceived message effectiveness would be higher for the health warning label, followed by the identity warning label, and then the control label.

The sample size was pre-determined by the parent study on nutrient labels. A post-hoc power analysis for this experiment indicated that the pre-determined sample size of 4,107 participants (∼1,370 per arm) would yield greater than 95% power to detect a minimum effect of Cohen’s *d*=.20 (a small effect size) when comparing each intervention arm to the control arm, assuming alpha=.05, correlation among repeated measures=.5, and 4 repeated measures.

Complete case analysis was used to handle any missing data (**Figure 1**). Of the 4,198 randomized participants, we excluded those who completed the survey implausibly quickly (defined as <1/3 of the median completion time) or completed less than 90% of the survey. We also checked for straight-lining behavior (responding to all the questions the same way, likely not truthfully), but none was detected and thus no exclusions made on this criterion. Finally, we excluded participants with responses (sessions) overlapping in time and dropped all but the first response for the remaining participants if they attempted or submitted multiple responses were excluded, resulting in an analytical sample of 4,107. All exclusions were pre-registered.

**Figure 1.**
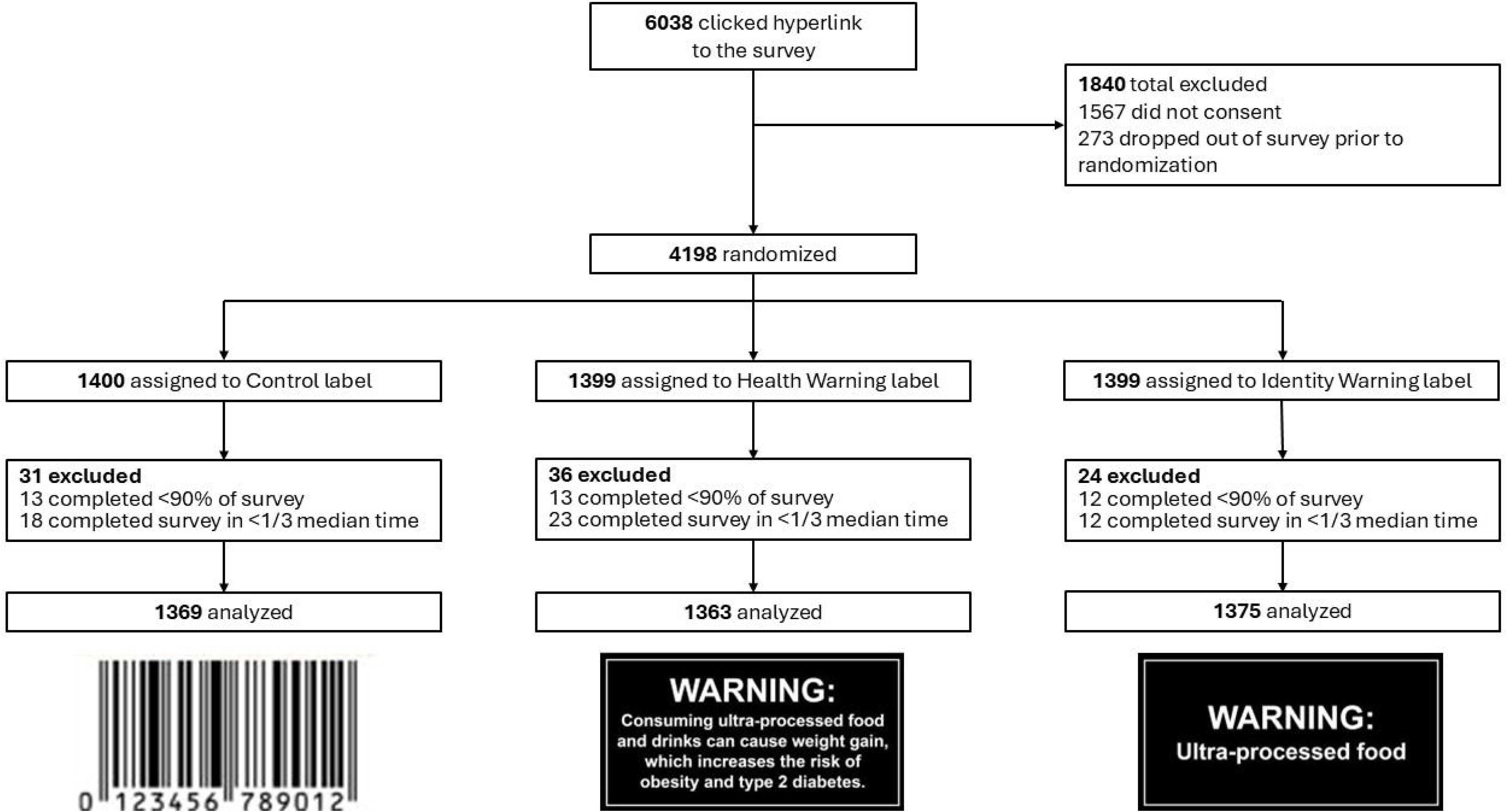
CONSORT diagram.

Initially, descriptive reports of unadjusted means or percentages for the primary and secondary outcomes were conducted. Then regression models were used to assess the effects of the labels –i.e., differences in the outcomes by treatment arm, comparing each of the two treatment arms (health warning label and identity warning label) to the control arm, as well as comparing the two treatment arms directly to each other. To assess effects on outcomes measured repeatedly across the different products, a mixed-effects logistic regression model for the dichotomous outcome (i.e., identification of products as ultra-processed) and mixed-effects linear regression models for the continuous outcomes (i.e., perceptions of healthfulness and purchase intentions) were used. These models treated the intercept as random in the regression models to account for repeated measures. All models included indicator variables for the labeling arm and for the product category. Alternatively, to assess effects on PME, which was assessed only once for each participant, we used a linear regression model. These models were used to estimate the average differential effects for all arms, representing the differences in predicted probabilities (for the dichotomous outcome) or means (for the continuous outcome) between label types. Analyses were conducted in Stata MP (v.18), with two-tailed tests, a critical alpha of 0.05, and 95% confidence intervals.

In separate models, exploratory analyses to examine whether educational attainment and English proficiency moderated the UPF labels’ effects on the primary outcome were conducted. Specifically, interaction terms between the labeling arms and education or English proficiency (specified as binary variables in separate models) were included and Wald chunk tests to determine the statistical significance of the joint interactions were used. These models were used to estimate the effects of the UPF labels for each level of the moderator.

Lastly, stratified analyses of the primary outcome using separate models for each category explored whether the impact of the warnings varied by product type.

## Results

Participants had a mean age of 35 years, and 50% identified as women (**Supplementary Table 1**). About a quarter (26%) of participants had a high school degree or less; a third (34%) had an associate degree, some college, or technical school; and 40% had a bachelor’s degree or higher. About half (49%) of participants reported limited English proficiency.

Identity warnings (70% correct) and health warnings (67% correct) both led to higher correct identification of UPF compared to control labels (54%, *p*<.001), with the identity warning having a larger impact than the health warning (*p*=.007) (**Figure 2**).

**Figure 2.**
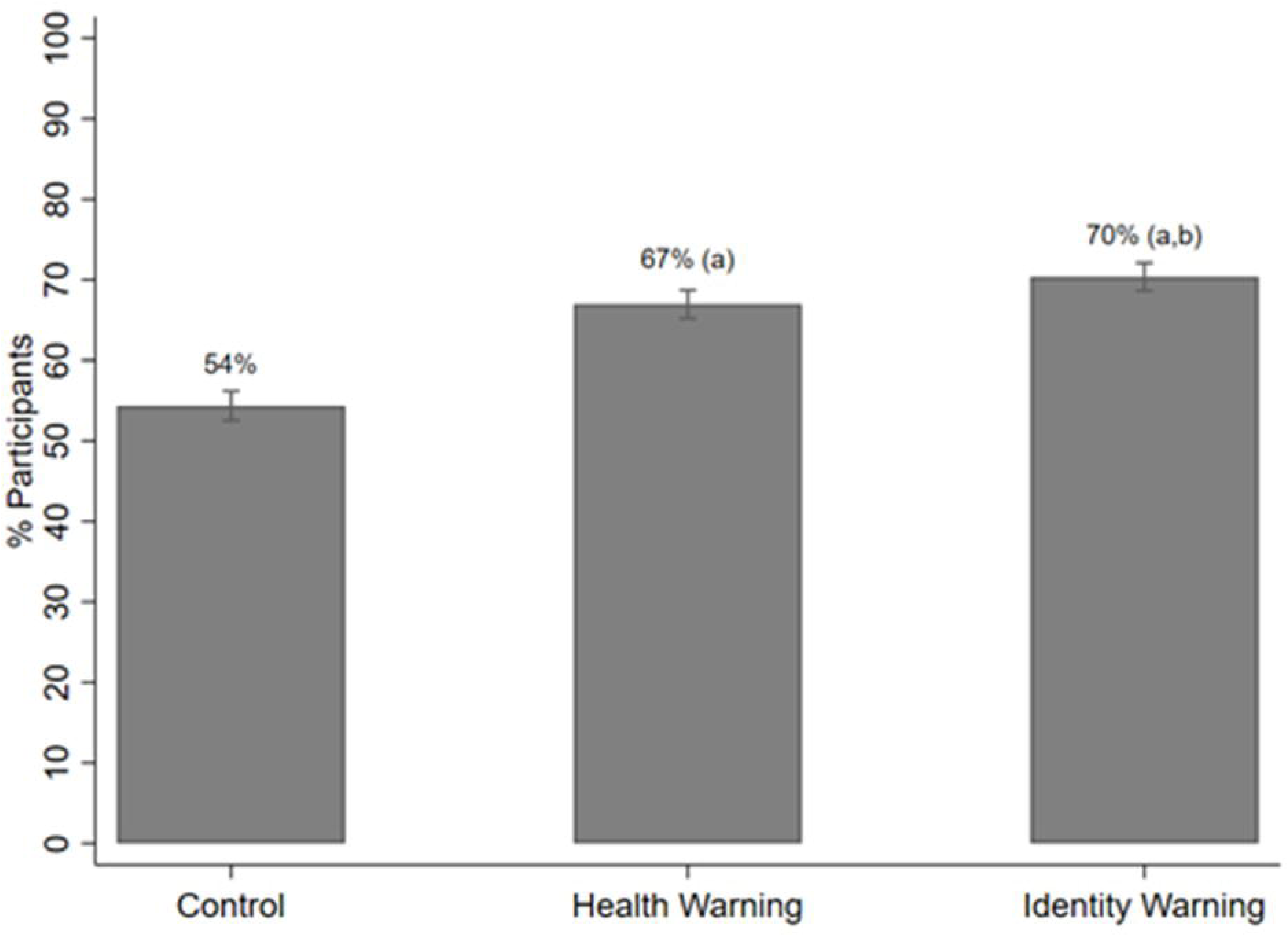
Percentage of participants who correctly identified the product as a UPF by trial arm. Note: *a* indicates a statistically significant difference (p < .001) in correct UPF identification between each treatment arm and the control group, and *b* indicates a statistically significant difference between the health warning and identity warning arms.

Compared to the control label, both the health warning and identity warning led to lower perceived healthfulness, reduced purchase intentions, and higher perceived message effectiveness for UPFs (**Table 1**) (*p*<0.001 for all comparisons). There were no differences in perceived healthfulness, purchase intentions, or perceived message effectiveness between the health warning and identity warning.

**Table 1.** Impact of UPF health warning and UPF identity warning labels on study outcomes.

|  | **Health Warning vs. Control** |  | **Identity Warning vs. Control** |  | **Identity Warning vs. Health Warning** |  |
| --- | --- | --- | --- | --- | --- | --- |
|  | Mean diff.<br>Percentage<br>points (pp)<br>(95% CI) | p-value | Mean diff.<br>pp<br>(95% CI) | p-value | Mean diff.<br>pp<br>(95% CI) | p-value |
| **Correct identification of UPF** | 13 pp<br>(10, 15) | <0.001 | 16 pp<br>(14, 19) | <0.001 | 3 pp<br>(1, 5) | 0.007 |
| **Perceived healthfulness** | -0.27<br>(-0.34, -0.21) | <0.001 | -0.33<br>(-0.39, -0.26) | <0.001 | -0.05<br>(-0.12, 0.02) | 0.166 |
| **Purchase intentions** | -0.26<br>(-0.34, -0.18) | <0.001 | -0.30<br>(-0.38, -0.22) | <0.001 | -0.04<br>(-0.12, 0.05) | 0.383 |
| **Perceived message effectiveness** | 0.88<br>(0.79, 0.97) | <0.001 | 0.85<br>(0.76, 0.94) | <0.001 | -0.03<br>(-0.11, 0.06) | 0.527 |

*Note.* The scale for the effects is 0-4.

Educational attainment and English proficiency moderated the effect of the labels on correct identification of UPFs (**Table 2**). Specifically, while the health warning improved participants’ ability to identify UPFs across all education and English proficiency levels, effects were greater for high vs. lower educated participants and for participants with limited vs. high English language proficiency.

**Table 2.**
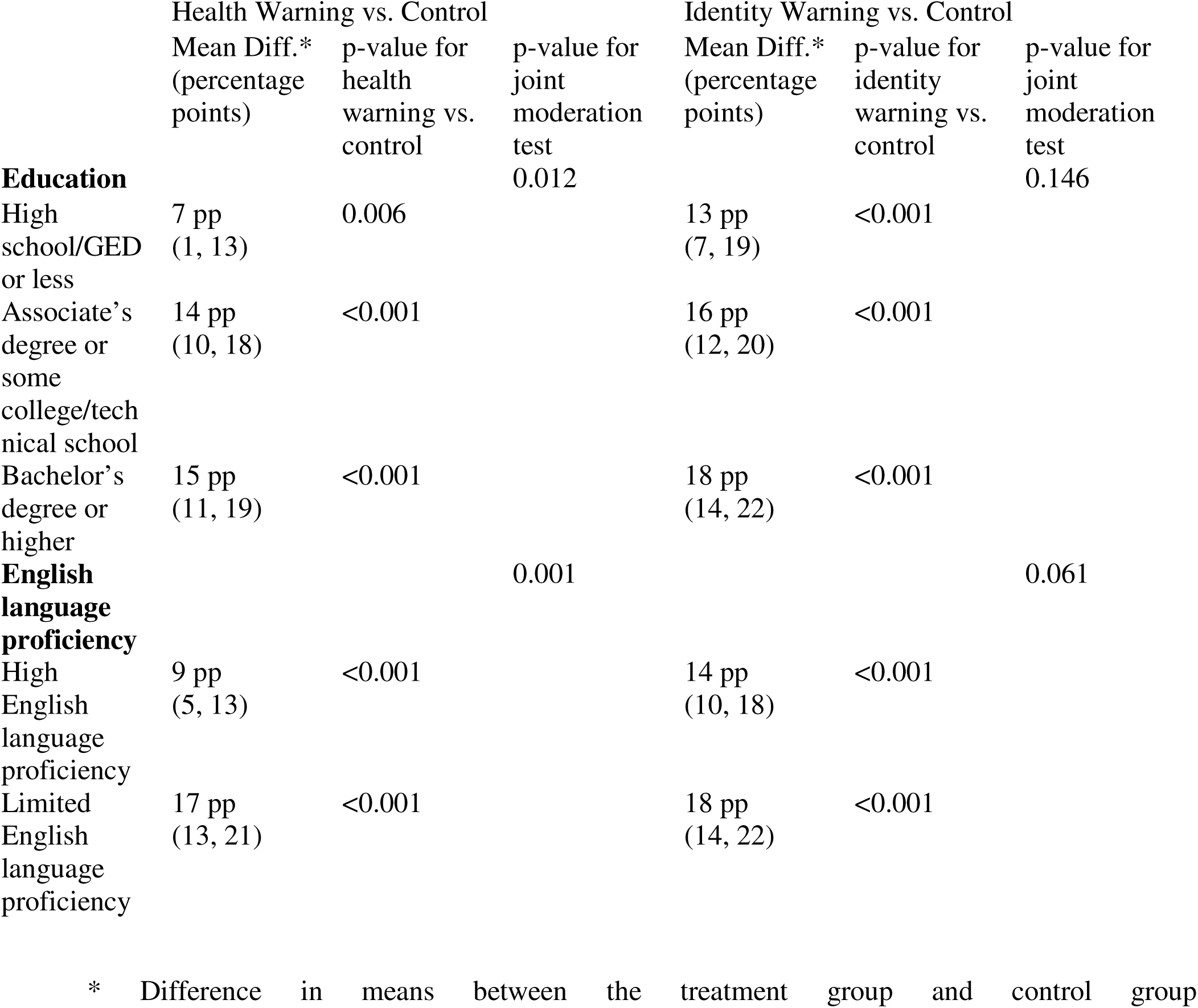
Moderation of education and English language proficiency on the impact of health warning and identity warning labels on correct identification of UPFs.

The overall direction and magnitude of results were similar for product types, with both warnings leading to greater correct identification of products as UPF, lower perceived healthfulness, and lower purchase intentions, compared to the control (**Supplementary Table 2**). Based on descriptive comparisons, the pattern of results across outcomes suggested the weakest effect for both warnings for pretzels and the strongest effect for yogurt.

## Discussion

In this online randomized trial of Latino adults in the US, front-of-package warnings improved participants’ ability to correctly identify products as ultra-processed. As predicted, both the UPF health warning (about the health effects of UPFs) and the UPF identity warning (stating that the product was a UPF) improved consumers’ ability to correctly identify products as ultra-processed when compared to the control, and the identity warning label led to increased ability to identify a product as ultra-processed compared to the health warning label. These results are in line with previous studies on UPF labels, which demonstrate that adding an “ultra-processed” label led to an increased ability to correctly identify products as UPFs compared to a control label.^25,47^

Both the identity warning and the health warning led to lower perceptions of the healthfulness of ultra-processed products, reduced intentions to purchase ultra-processed products, and higher perceived effectiveness of the label at discouraging consumption of ultra-processed products when compared to the control. Previous research on UPF labels has found similar results, with UPF labels eliciting higher PME and more thinking about the health risks of UPF consumption.^48^ In this study, both the identify warning and the health warning led to similar changes in perceived healthfulness, purchase intentions, and PME as one another. Prior studies of health and identity warnings about nutrients have found both similar and conflicting results. For example, our results contrast with a previous meta-analysis and randomized experiment, both of which found that health warnings led to greater reductions in hypothetical purchases and higher PME compared to identity warnings.^19,31^ However, other studies assessing reactions to sugary drinks have found different results. In one randomized experiment of U.S. adults in which approximately half of the sample identified as Latino, nutrient and health warnings did not differ from each other on perceptions of healthfulness, intentions to purchase, or PME.^43^ Additionally, in a randomized controlled trial in Singapore in which participants were asked to shop in an experimental online grocery store, hypothetical purchases of high-in-sugar products were similar in the identity warning and health warning arms.^32^ Differences between this study and prior research may reflect that prior studies examined identity warnings about nutrients, rather than processing level. Additional research is needed to understand when health warnings are likely to outperform identity warnings, including UPFs.

The health warning tested in this study described multiple health effects (weight gain, obesity, and type 2 diabetes). Some previous research indicates that warnings with multiple health effects may be more effective than warnings about one health effect.^49^ However, other studies support the theory that “less is more” when presenting consumers with information about quality.^50^ Future research could explore the “sweet spot” for the number of health effects displayed on a health warning for the UPF, as well as whether certain health effects elicit greater consumer response than others.

English language proficiency and education level moderated the impact of warning type on correct UPF identification. This pattern appears to be driven by the health warning having a greater impact on correct identification of UPFs for those with higher education (vs. lower education) and among those with limited English language proficiency (vs. higher English proficiency). These results were somewhat surprising: if participants with lower education were less responsive to the health warning, perhaps due to lower comprehension of the label, one might assume that participants with limited English proficiency might also be less responsive to the health warning for similar reasons. One possible explanation for the discrepancy is that participants with limited English proficiency had higher education levels compared to those with high English proficiency in the sample. The overlap among these two demographics might explain the greater impact of the health warning among both participants with limited English proficiency and with higher education. Another possible explanation could be that participants with limited English proficiency took more time to read the label, leading to its greater effectiveness in this group. That explanation is consistent with previous research finding that participants with limited English proficiency tend to perceive health warnings as more effective than their higher-English-proficiency counterparts^43,51^ and were more behaviorally responsive to English nutrient warning labels.^52^ More research will be needed to understand the psychological mechanisms underlying how warnings work in different populations and to design warnings that are effective for all consumer types.

### Limitations

Strengths of the study include the high number of participants (nearly half of the sample) with limited English proficiency, the use of evidence-based stimuli and multiple types of real products, and the comparison between UPF health warnings and UPF identity warnings, which has not been previously examined. One limitation is that participants were only exposed to the labels on four products. Additionally, participants were exposed to labels for a short period of time in an online setting with hypothetical outcomes, which limits the generalizability of the findings to other settings and products and precluded the ability to measure behavioral outcomes. Future research should be conducted in real-world food purchasing settings with additional products to understand whether and how UPF warning labels impact real-world behaviors. Understanding how UPF warnings affect behavior and beliefs in a real-world context is especially important given the high proportion of packaged foods that would receive a UPF warning as well as the growing interest in regulating UPFs among American consumers and policymakers.

## Conclusions

In a randomized experiment, UPF health warnings and identity warnings improved participants’ ability to identify ultra-processed products and reduced perceived healthfulness and intentions to purchase UPFs. While identity warning labels increased participant’s ability to correctly identify ultra-processed products compared to health warning labels, there were no significant differences between the two label types’ influence on healthfulness perceptions or purchase intentions. The impact of health warnings (vs. control warnings) was stronger for those with limited English proficiency and higher levels of education than those with higher English proficiency and lower levels of education, respectively. These results suggest the promise of UPF warnings — including health and identity warnings — for informing consumers and encouraging healthier purchases.

## Supporting information

Highlights

Declaration of Interest Statement

CONSORT Checklist

Credit Statement

## Acknowledgements

We thank Emily Busey for designing the study stimuli and Callie Whitesell and Angela Viviana Martinez for programming the online survey.

## Data Availability Statement

The data that support the findings of this study are openly available in the UNC Dataverse at https://dataverse.unc.edu/dataset.xhtml?persistentId=doi:10.15139/S3/VRUWYO.

## References

1. Monteiro CA, Cannon G, Levy RB, et al. Ultra-processed foods: what they are and how to identify them. Public Health Nutrition. 2019;22(5):936–941. doi:10.1017/S1368980018003762

2. Williams A, Couch C, Emmerich S, Ogburn D. Ultra-processed food consumption among youth and adults: United States, August *2021–August* 2023. Vol. 536. 2025:1–11. *NCHS Data Brief*.

3. Moradi S, Entezari MH, Mohammadi H, et al. Ultra-processed food consumption and adult obesity risk: a systematic review and dose-response meta-analysis. Crit Rev Food Sci Nutr. 2023;63(2):249–260. doi:10.1080/10408398.2021.1946005

4. Vitale M, Costabile G, Testa R, et al. Ultra-Processed Foods and Human Health: A Systematic Review and Meta-Analysis of Prospective Cohort Studies. Adv Nutr. Jan 2024;15(1):100121. doi:10.1016/j.advnut.2023.09.009

5. Pagliai G, Dinu M, Madarena MP, Bonaccio M, Iacoviello L, Sofi F. Consumption of ultra-processed foods and health status: a systematic review and meta-analysis. Br J Nutr. Feb 14 2021;125(3):308–318. doi:10.1017/s0007114520002688

6. Delpino FM, Figueiredo LM, Bielemann RM, et al. Ultra-processed food and risk of type 2 diabetes: a systematic review and meta-analysis of longitudinal studies. Int J Epidemiol. Aug 10 2022;51(4):1120–1141. doi:10.1093/ije/dyab247

7. Guo L, Li F, Tang G, et al. Association of ultra-processed foods consumption with risk of cardio-cerebrovascular disease: A systematic review and meta-analysis of cohort studies. Nutr Metab Cardiovasc Dis. Nov 2023;33(11):2076–2088. doi:10.1016/j.numecd.2023.07.005

8. Lane MM, Gamage E, Du S, et al. Ultra-processed food exposure and adverse health outcomes: umbrella review of epidemiological meta-analyses. BMJ. 2024;384:e077310. doi:10.1136/bmj-2023-077310

9. Taneri PE, Wehrli F, Roa-Díaz ZM, et al. Association Between Ultra-Processed Food Intake and All-Cause Mortality: A Systematic Review and Meta-Analysis. American Journal of Epidemiology. 2022;191(7):1323–1335. doi:10.1093/aje/kwac039

10. Hall KD, Ayuketah A, Brychta R, et al. Ultra-Processed Diets Cause Excess Calorie Intake and Weight Gain: An Inpatient Randomized Controlled Trial of Ad Libitum Food Intake. Cell Metab. Jul 2 2019;30(1):67–77.e3. doi:10.1016/j.cmet.2019.05.008

11. Hamano S, Sawada M, Aihara M, et al. Ultra-processed foods cause weight gain and increased energy intake associated with reduced chewing frequency: A randomized, open-label, crossover study. Diabetes Obes Metab. Nov 2024;26(11):5431–5443. doi:10.1111/dom.15922

12. Taillie LS, Hall MG, Popkin BM, Ng SW, Murukutla N. Experimental Studies of Front-of-Package Nutrient Warning Labels on Sugar-Sweetened Beverages and Ultra-Processed Foods: A Scoping Review. Nutrients. Feb 22 2020;12(2)doi:10.3390/nu12020569

13. Scapin T, Fernandes AC, Curioni CC, et al. Influence of sugar label formats on consumer understanding and amount of sugar in food choices: a systematic review and meta-analyses. Nutrition Reviews. 2020;79(7):788–801. doi:10.1093/nutrit/nuaa108

14. Bopape M, De Man J, Taillie LS, Ng SW, Murukutla N, Swart R. Effect of different front-of-package food labels on identification of unhealthy products and intention to purchase the products- A randomised controlled trial in South Africa. Appetite. Dec 1 2022;179:106283. doi:10.1016/j.appet.2022.106283

15. Mora-Plazas M, Higgins ICA, Gomez LF, et al. Impact of nutrient warning labels on Colombian consumers’ selection and identification of food and drinks high in sugar, sodium, and saturated fat: A randomized controlled trial. PLoS One. 2024;19(6):e0303514. doi:10.1371/journal.pone.0303514

16. Temple NJ. Front-of-package food labels: A narrative review. Appetite. 2020/01/01/ 2020;144:104485. 10.1016/j.appet.2019.104485

17. Song J, Brown MK, Tan M, et al. Impact of color-coded and warning nutrition labelling schemes: A systematic review and network meta-analysis. PLoS medicine. 2021;18(10):e1003765.

18. An R, Liu J, Liu R, Barker AR, Figueroa RB, McBride TD. Impact of Sugar-Sweetened Beverage Warning Labels on Consumer Behaviors: A Systematic Review and Meta-Analysis. American Journal of Preventive Medicine. 2021/01/01/ 2021;60(1):115–126. 10.1016/j.amepre.2020.07.003

19. Grummon AH, Hall MG. Sugary drink warnings: A meta-analysis of experimental studies. PLoS medicine. May 2020;17(5):e1003120. doi:10.1371/journal.pmed.1003120

20. Taillie LS, Higgins ICA, Lazard AJ, Miles DR, Blitstein JL, Hall MG. Do sugar warning labels influence parents’ selection of a labeled snack for their children? A randomized trial in a virtual convenience store. Appetite. 2022/08/01/ 2022;175:106059. 10.1016/j.appet.2022.106059

21. Ikonen I, Sotgiu F, Aydinli A, Verlegh PW. Consumer effects of front-of-package nutrition labeling: An interdisciplinary meta-analysis. Journal of the Academy of Marketing Science. 2020;48(3):360–383.

22. Hall MG, Grummon AH, Higgins ICA, et al. The impact of pictorial health warnings on purchases of sugary drinks for children: A randomized controlled trial. PLOS Medicine. 2022;19(2):e1003885. doi:10.1371/journal.pmed.1003885

23. Sarmiento-Santos J, Souza MBN, Araujo LS, Pion JMV, Carvalho RA, Vanin FM. Consumers’ Understanding of Ultra-Processed Foods. Foods. May 7 2022;11(9)doi:10.3390/foods11091359

24. Machín L, Antúnez L, Curutchet MR, Ares G. The heuristics that guide healthiness perception of ultra-processed foods: a qualitative exploration. Public Health Nutr. Nov 2020;23(16):2932–2940. doi:10.1017/s1368980020003158

25. D’Angelo Campos A, Ng SW, Duran AC, et al. “Warning: ultra-processed“: an online experiment examining the impact of ultra-processed warning labels on consumers’ product perceptions and behavioral intentions. Int J Behav Nutr Phys Act. Oct 9 2024;21(1):115. doi:10.1186/s12966-024-01664-w

26. Pedro-Botet L, Muns MD, Solà R, et al. Level of understanding and consumption of ultra-processed food in a Mediterranean population: A cross-sectional study. Nutr Metab Cardiovasc Dis. Apr 2022;32(4):889–896. doi:10.1016/j.numecd.2021.11.002

27. Bhawra J, Kirkpatrick SI, Hall MG, Vanderlee L, White CM, Hammond D. Patterns and correlates of nutrition knowledge across five countries in the 2018 international food policy study. Nutr J. Mar 16 2023;22(1):19. doi:10.1186/s12937-023-00844-x

28. Ares G, Vidal L, Allegue G, et al. Consumers’ conceptualization of ultra-processed foods. Appetite. Oct 1 2016;105:611–7. doi:10.1016/j.appet.2016.06.028

29. Myers I. Interactive map: Tracking state food chemical regulation in the U.S. Environmental Working Group. Accessed November 17, 2025. https://www.ewg.org/news-insights/news/2025/10/interactive-map-tracking-state-food-chemical-regulation-us

30. Childhood Diabetes Reduction Act of 2024, S.4195, 118th Congress, 2nd sess (Sanders B 2024). April 18. https://www.congress.gov/bill/118th-congress/senate-bill/4195/text

31. Grummon AH, Hall MG, Taillie LS, Brewer NT. How should sugar-sweetened beverage health warnings be designed? A randomized experiment. Prev Med. Apr 2019;121:158–166. doi:10.1016/j.ypmed.2019.02.010

32. Ang FJL, Agrawal S, Finkelstein EA. Pilot randomized controlled trial testing the influence of front-of-pack sugar warning labels on food demand. BMC Public Health. Feb 7 2019;19(1):164. doi:10.1186/s12889-019-6496-8

33. 2020 Census Statistics Highlight Local Population Changes and Nation’s Racial and Ethnic Diversity. United States Census Bureau; August 12, 2021.

34. Fryar CD, Carroll MD, Ogden CL. Prevalence of overweight, obesity, and severe obesity among adults aged 20 and over: United States, 1960–1962 through 2015–2016. 2018;NCHS Health E-Stats.

35. Guerra ZC, Moore JR, Londoño T, Castro Y. Associations of Acculturation and Gender with Obesity and Physical Activity among Latinos. Am J Health Behav. Jun 23 2022;46(3):324–336. doi:10.5993/ajhb.46.3.11

36. Aguayo-Mazzucato C, Diaque P, Hernandez S, Rosas S, Kostic A, Caballero AE. Understanding the growing epidemic of type 2 diabetes in the Hispanic population living in the United States. Diabetes Metab Res Rev. Feb 2019;35(2):e3097. doi:10.1002/dmrr.3097

37. National diabetes statistics report, 2020 : estimates of diabetes and its burden in the United States. 14/20

38. Herbert BM, Johnson AE, Paasche-Orlow MK, Brooks MM, Magnani JW. Disparities in Reporting a History of Cardiovascular Disease Among Adults With Limited English Proficiency and Angina. JAMA Netw Open. Dec 1 2021;4(12):e2138780. doi:10.1001/jamanetworkopen.2021.38780

39. Kim EJ, Kim T, Paasche-Orlow MK, Rose AJ, Hanchate AD. Disparities in Hypertension Associated with Limited English Proficiency. J Gen Intern Med. Jun 2017;32(6):632–639. doi:10.1007/s11606-017-3999-9

40. Hall MG, Lee CJY, Campos AD, et al. Effects of front-of-package nutrition labels in Latine and limited English proficiency populations: A randomized trial. medRxiv. May 11 2025;doi:10.1101/2025.05.09.25327177

41. Dietrich S, Hernandez E. Language Use in the United States: 2019. 2022. https://www.census.gov/content/dam/Census/library/publications/2022/acs/acs-50.pdf

42. Hall MG, Grummon AH, Maynard OM, Kameny MR, Jenson D, Popkin BM. Causal Language in Health Warning Labels and US Adults’ Perception: A Randomized Experiment. American Journal of Public Health. 2019;109(10):1429–1433. doi:10.2105/ajph.2019.305222

43. Hall MG, Lazard AJ, Grummon AH, et al. Designing warnings for sugary drinks: A randomized experiment with Latino parents and non-Latino parents. Preventive medicine. Jul 2021;148:106562. doi:10.1016/j.ypmed.2021.106562

44. Grummon AH, Taillie LS, Golden SD, Hall MG, Ranney LM, Brewer NT. Sugar-Sweetened Beverage Health Warnings and Purchases: A Randomized Controlled Trial. Am J Prev Med. Nov 2019;57(5):601–610. doi:10.1016/j.amepre.2019.06.019

45. Bollard T, Maubach N, Walker N, Ni Mhurchu C. Effects of plain packaging, warning labels, and taxes on young people’s predicted sugar-sweetened beverage preferences: an experimental study. Int J Behav Nutr Phys Act. Sep 1 2016;13(1):95. doi:10.1186/s12966-016-0421-7

46. Baig SA, Noar SM, Gottfredson NC, Boynton MH, Ribisl KM, Brewer NT. UNC Perceived Message Effectiveness: Validation of a Brief Scale. Ann Behav Med. Jul 17 2019;53(8):732–742. doi:10.1093/abm/kay080

47. Srour B, Hercberg S, Galan P, et al. Effect of a new graphically modified Nutri-Score on the objective understanding of foods’ nutrient profile and ultraprocessing: a randomised controlled trial. BMJ Nutr Prev Health. Jun 2023;6(1):108–118. doi:10.1136/bmjnph-2022-000599

48. D’Angelo Campos A, Ng SW, McNeel K, Hall MG. How Promising Are “Ultraprocessed” Front-of-Package Labels? A Formative Study with US Adults. Nutrients. Apr 6 2024;16(7)doi:10.3390/nu16071072

49. Noar SM, Kelley DE, Boynton MH, et al. Identifying principles for effective messages about chemicals in cigarette smoke. Preventive medicine. Jan 2018;106:31–37. doi:10.1016/j.ypmed.2017.09.005

50. Peters E, Dieckmann N, Dixon A, Hibbard JH, Mertz CK. Less Is More in Presenting Quality Information to Consumers. Medical Care Research and Review. 2007;64(2):169–190. doi:10.1177/10775587070640020301

51. Hall MG, Grummon AH, Whitesell C, et al. Evaluating text, icon, and graphic nutrition labels: An eye tracking experiment with Latino adults in the US. Appetite. Jan 1 2025;204:107745. doi:10.1016/j.appet.2024.107745

