## Supplementary material for "Impacts of warning labels on ultra-processed foods among Latino adults: A randomized trial": Highlights

- Ultra-processed foods are linked to adverse health outcomes
- Front-of-package warning labels on ultra-processed products help identify them
- Identity and health warning labels on ultra-processed foods discourage consumption
- English proficiency and education can impact the efficacy of health warning labels
