## Supplementary material for "Impacts of warning labels on ultra-processed foods among Latino adults: A randomized trial": Declaration of Interest Statement

**Conflict of interest statement:**
This research was supported by Healthy Eating Research, a national program of the Robert Wood Johnson Foundation. J.F. was supported by USDA National Institute of Food and Agriculture (NIFA) Hatch project #7005204. A.H.G. was supported by K01 HL158608 from the National Institutes of Health. C.E.P received support from the Population Research Training grant (T32 HD007168) and the Population Research Infrastructure Program (P2C HD050924) awarded to the Carolina Population Center at the University of North Carolina at Chapel Hill by the Eunice Kennedy Shriver National Institute of Child Health and Human Development. A.A.M. received support from Bloomberg Philanthropies. The other authors did not have any funding sources. The funders did not play a role in the study design; collection, analysis, and interpretation of data; writing the report; or the decision to submit the paper.

**Financial disclosure**

No financial disclosures were reported by the authors of this paper.
