## Supplementary Materials for "Impacts of warning labels on ultra-processed foods among Latino adults: A randomized trial"

**Supplementary Materials for “Impacts of Warning Labels on Ultra-Processed Foods - A Randomized Controlled Trial”**

### **Supplementary Table 1. Sociodemographic characteristics of the sample (n = 4,107).**

|  | **Control**  **(*n* = 1,369)** | **Health Warning**  **(*n* = 1,363)** | **Identity Warning (*n* = 1,375)** | **Total**  **(*n* = 4,107)** |
| --- | --- | --- | --- | --- |
|  | **n (%)** | **n (%)** | **n (%)** | **n (%)** |
| **Age** |  |  |  |  |
| 18-25 years | 268 (20%) | 312 (23%) | 274 (20%) | 854 (21%) |
| 26-35 years | 456 (33%) | 415 (30%) | 442 (32%) | 1,313 (32%) |
| 36-45 years | 473 (35%) | 466 (34%) | 480 (35%) | 1,419 (35%) |
| 46-55 years | 172 (13%) | 170 (12%) | 179 (13%) | 521 (13%) |
| **Gender** |  |  |  |  |
| Woman | 668 (49%) | 684 (50%) | 687 (50%) | 2,039 (50%) |
| Man | 668 (49%) | 658 (48%) | 664 (48%) | 1,990 (48%) |
| Non-binary or self-described | 31 (2%) | 21 (2%) | 22 (2%) | 74 (2%) |
| **Annual household income** |  |  |  |  |
| $14,999 or less | 193 (14%) | 214 (16%) | 201 (15%) | 608 (15%) |
| $15,000 to $34,999 | 219 (16%) | 205 (15%) | 200 (15%) | 624 (15%) |
| $35,000 to $74,999 | 262 (19%) | 236 (17%) | 281 (20%) | 779 (19%) |
| $75,000 – $149,999 | 427 (31%) | 438 (32%) | 416 (30%) | 1,281 (31%) |
| $150,000 or more | 242 (18%) | 253 (19%) | 257 (19%) | 752 (18%) |
| **English language proficiency** |  |  |  |  |
| Limited | 671 (49%) | 668 (49%) | 689 (50%) | 2028 (49%) |
| High | 698 (51%) | 695 (51%) | 686 (50%) | 2,079 (51%) |
| **Education** |  |  |  |  |
| High school degree/GED or less | 345 (25%) | 355 (26%) | 350 (25%) | 1050 (26%) |
| Associate’s degree or some college/technical school | 462 (34%) | 465 (34%) | 474 (35%) | 1401 (34%) |
| Bachelor’s degree or higher | 560 (41%) | 543 (40%) | 549 (40%) | 1652 (40%) |
| **Race** |  |  |  |  |
| White | 766 (56%) | 786 (58%) | 772 (56%) | 2324 (57%) |
| Black or African American | 227 (17%) | 220 (16%) | 222 (16%) | 669 (16%) |
| Indigenous or American Indian | 137 (10%) | 130 (10%) | 151 (11%) | 418 (10%) |
| Other | 233 (17%) | 222 (16%) | 227 (17%) | 682 (17%) |
| **Heritage** |  |  |  |  |
| Mexican | 447 (33%) | 454 (33%) | 454 (33%) | 1355 (33%) |
| Spanish/Spainard | 239 (17%) | 208 (15%) | 236 (17%) | 683 (17%) |
| Puerto Rican | 143 (10%) | 151 (11%) | 136 (10%) | 430 (10%) |
| Cuban | 130 (9%) | 135 (10%) | 107 (8%) | 372 (9%) |
| Venezuelan | 59 (4%) | 72 (5%) | 86 (6%) | 217 (5%) |
| Dominican | 55 (4%) | 55 (4%) | 57 (4%) | 167 (4%) |
| Colombian | 46 (3%) | 51 (4%) | 66 (5%) | 163 (4%) |
| Honduran | 40 (3%) | 40 (3%) | 49 (4%) | 129 (3%) |
| Salvadoran | 39 (3%) | 44 (3%) | 43 (3%) | 126 (3%) |
| Ecuadorian | 35 (3%) | 35 (3%) | 34 (2%) | 104 (3%) |
| Guatemalan | 43 (3%) | 30 (2%) | 26 (2%) | 99 (2%) |
| Peruvian | 21 (2%) | 28 (2%) | 21 (2%) | 70 (2%) |
| Brazilian | 14 (1%) | 20 (1%) | 18 (1%) | 52 (1%) |
| Other | 56 (4%) | 40 (3%) | 39 (3%) | 135 (3%) |

*Note.* Percentage of missing demographic data ranged from 0 to 3%.

### **Supplementary Table 2. Impact of UPF health warning and UPF identity warning labels on study outcomes stratified by product type (n = 4,107, n observations = 16,416).**

|  | **Health Warning vs. Control** | | **Identity Warning vs. Control** | |
| --- | --- | --- | --- | --- |
| Product Type | Mean diff. (95% CI) | p-value | Mean diff. (95% CI) | p-value |
| **Cereal** |  |  |  |  |
| Correct UPF identification | 12 pp  (8, 16) | <0.001 | 15 pp  (11, 18) | <0.001 |
| Perceived healthfulness | -0.27  (-0.35, -0.19) | <0.001 | -0.35  (-0.44, -0.27) | <0.001 |
| Purchase intentions | -0.23  (-0.33, -0.14) | <0.001 | -0.30  (-0.40, -0.20) | <0.001 |
| **Drink** |  |  |  |  |
| Correct UPF identification | 11 pp  (7, 14) | <0.001 | 13 pp  (10, 17) | <0.001 |
| Perceived healthfulness | -0.28  (-0.36, -0.19) | <0.001 | -0.32  (-0.40, -0.22) | <0.001 |
| Purchase intentions | -0.27 (-0.37, -0.16) | <0.001 | -0.27  (-0.38, -0.17) | <0.001 |
| **Pretzel** |  |  |  |  |
| Correct UPF identification | 10 pp  (7, 14) | <0.001 | 15 pp  (11, 18) | <0.001 |
| Perceived healthfulness | -0.17  (-0.26, -0.09) | <0.001 | -0.24  (-0.33, -0.16) | <0.001 |
| Purchase intentions | -0.20  (-0.30, -0.10) | <0.001 | -0.23  (-0.33, -0.13) | <0.001 |
| **Yogurt** |  |  |  |  |
| Correct UPF identification | 17 pp  (13, 20) | <0.001 | 20 pp  (17, 24) | <0.001 |
| Perceived healthfulness | -0.38  (-0.46, -0.30) | <0.001 | -0.39  (-0.47, -0.31) | <0.001 |
| Purchase intentions | -0.34  (-0.44, -0.25) | <0.001 | -0.38  (-0.48, -0.28) | <0.001 |

### **Supplementary Figure 1. Stimuli used in experiment.**

| Control | Identity Warning | Health Warning |
| --- | --- | --- |
| 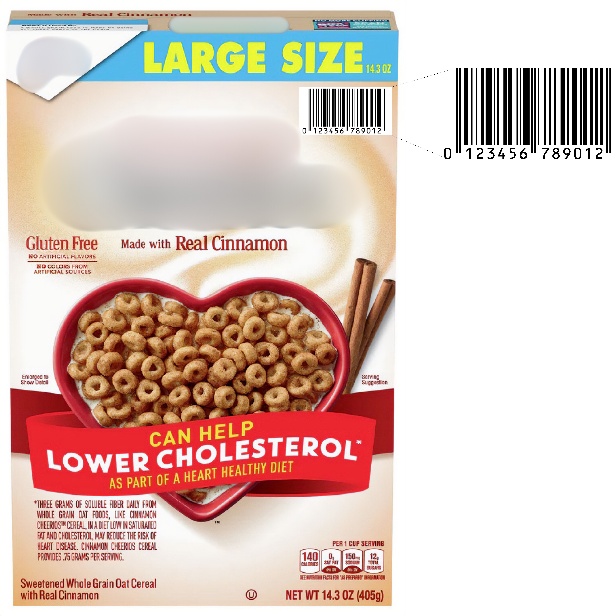 | 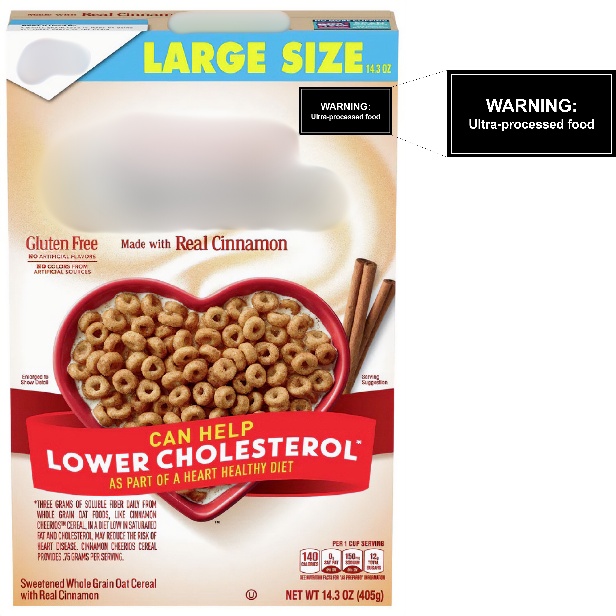 | 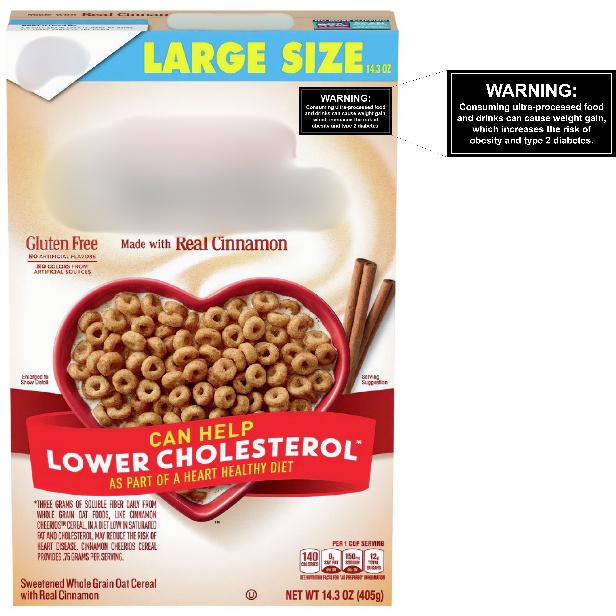 |
| 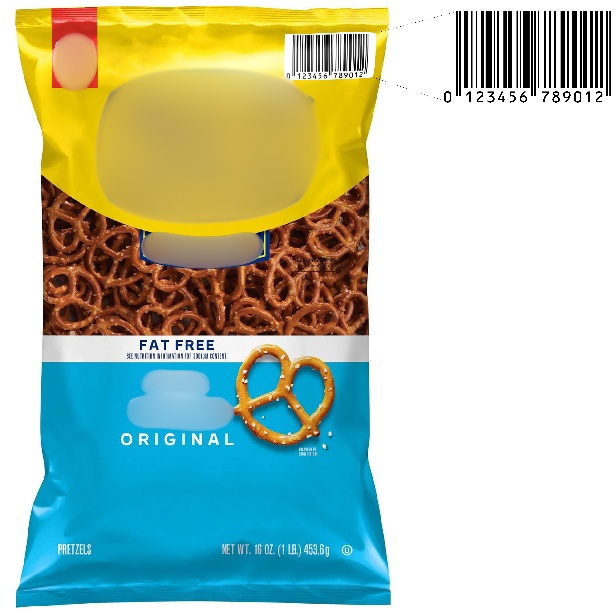 | 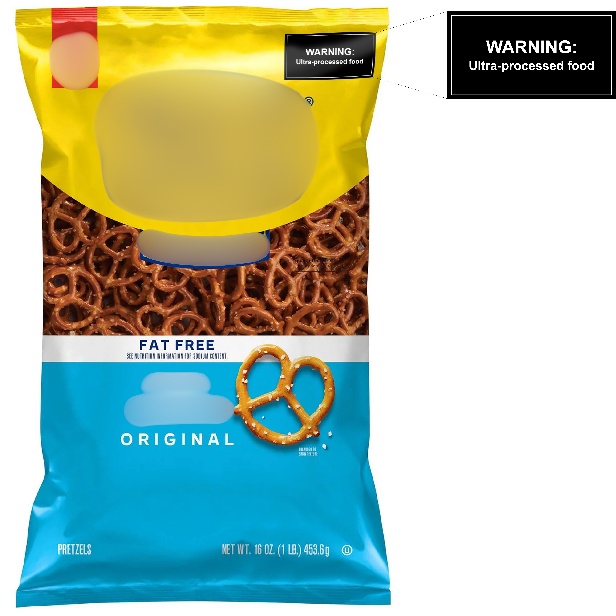 | 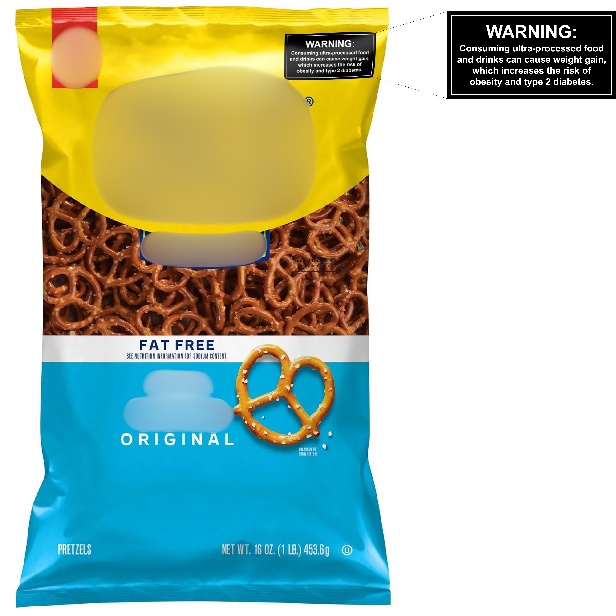 |
| 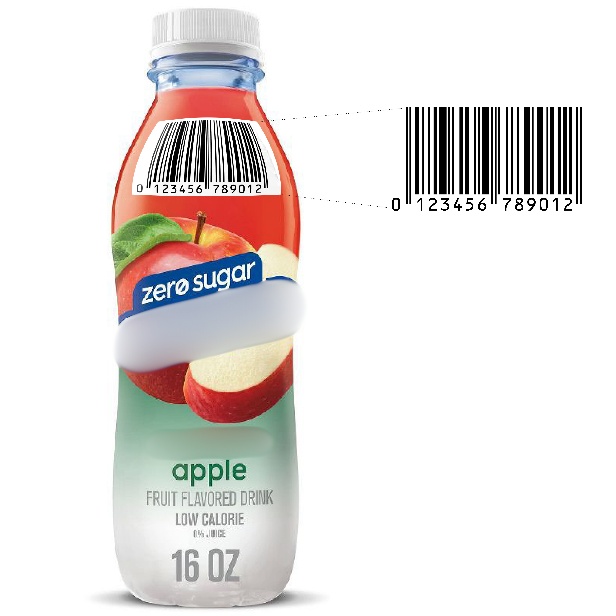 | 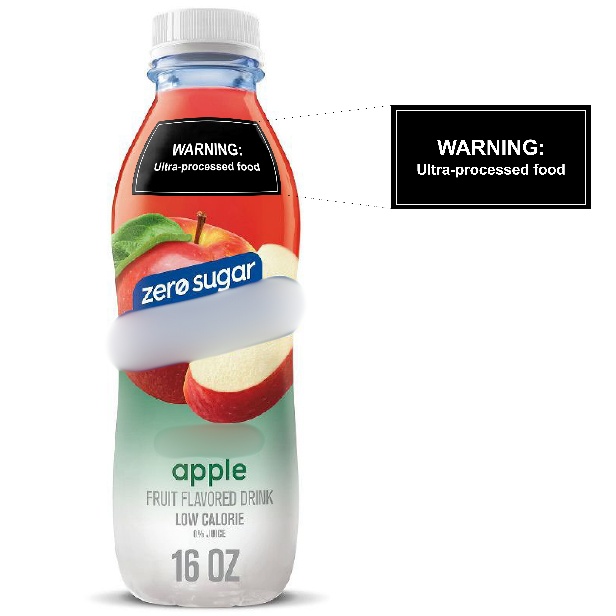 | 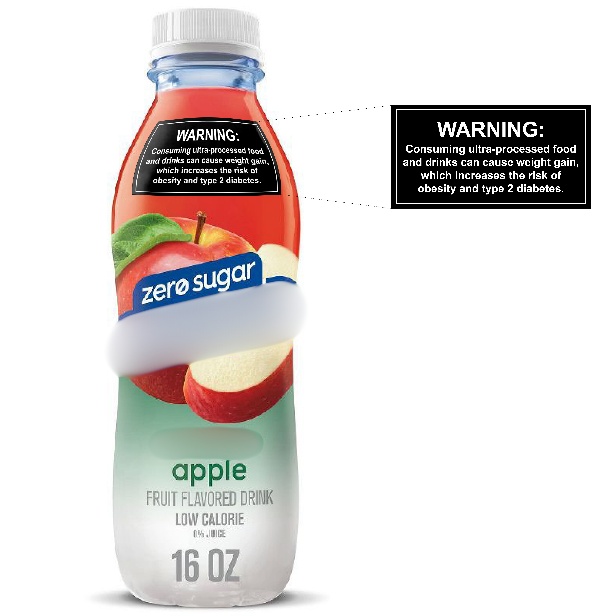 |
| 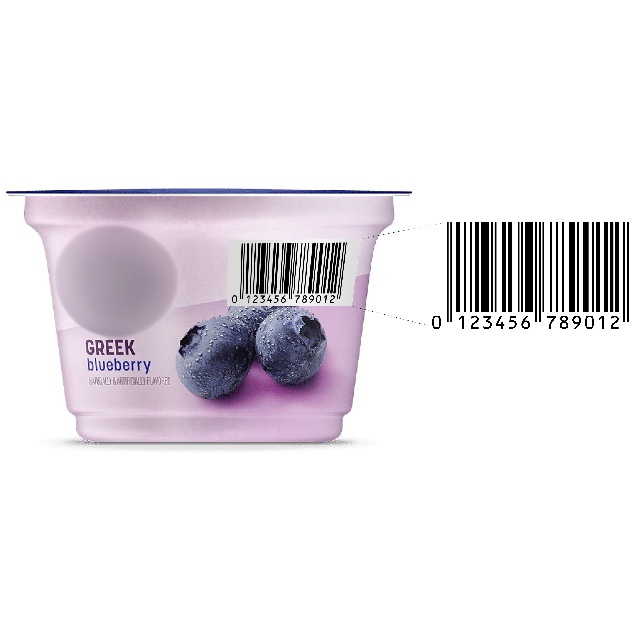 | 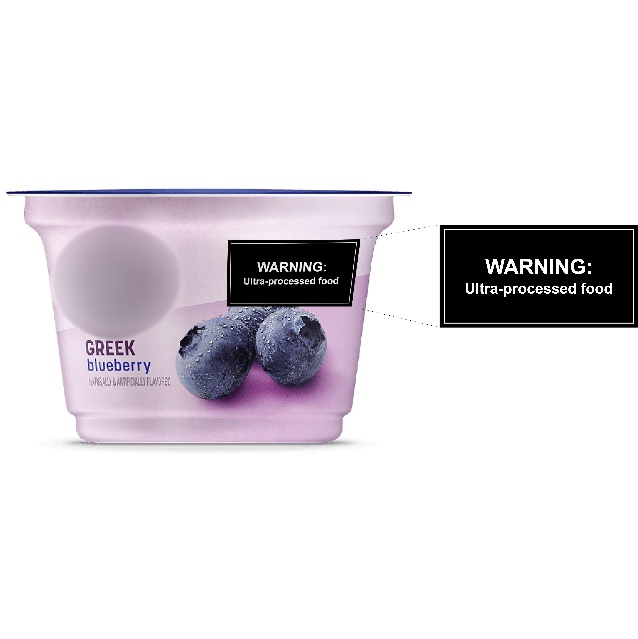 | 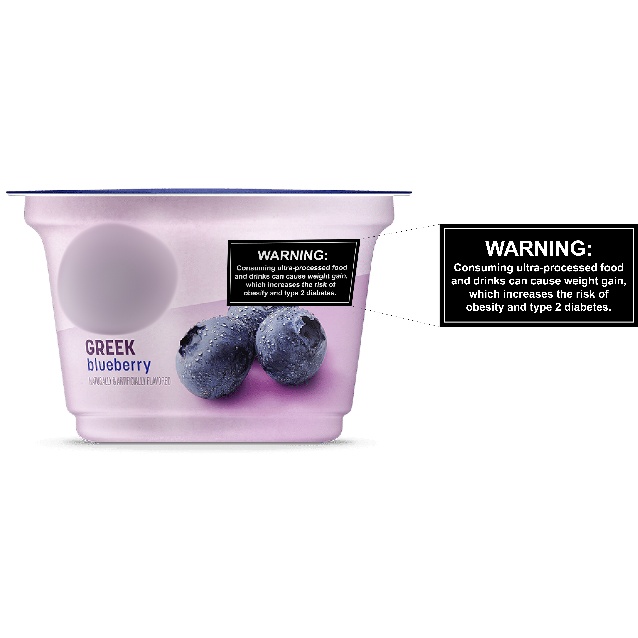 |

Note: Branding was not obscured during the experiment

### **Supplementary Figure 2. Warning labels used in experiment.**

| Control | Identity Warning | Health Warning |
| --- | --- | --- |
| 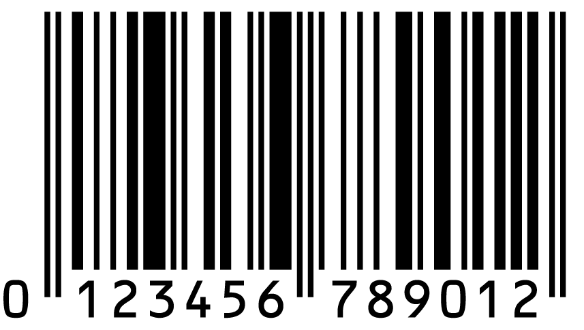 | 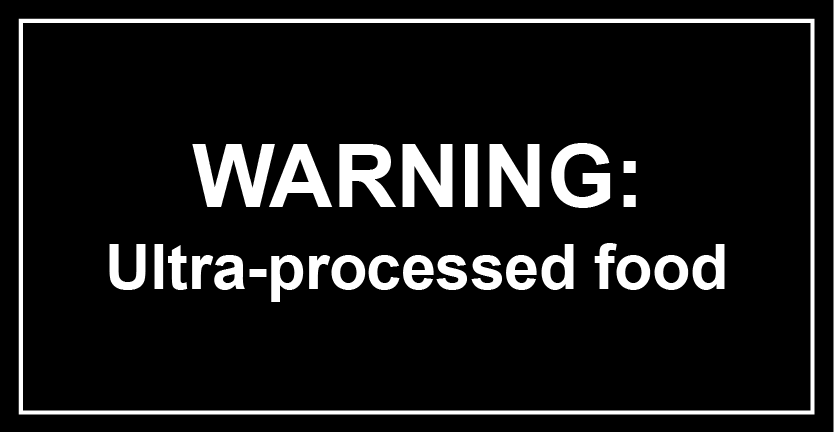 | 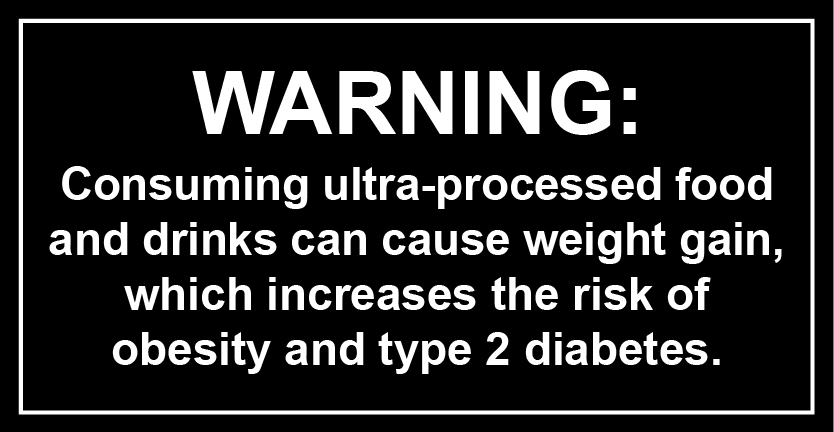 |
