## Supplementary material for "Impacts of warning labels on ultra-processed foods among Latino adults: A randomized trial": Credit Statement

**Author contributions: Lindsey Smith Taillie:** Conceptualization, Methodology, Supervision, Writing- Original draft preparation. **Violet Noe:** Methodology, Project Administration, Writing- Original draft preparation. **Mrignyani Sehgal:** Formal analysis, Visualization, Writing- Original draft preparation. **Aline D’Angelo Campos:** Conceptualization, Methodology, Writing- Reviewing and Editing. **Anna Grummon:** Conceptualization, Methodology, Writing- Reviewing and Editing, Funding acquisition. **Jennifer Falbe:** Conceptualization, Methodology, Writing- Reviewing and Editing. **Aviva Musicus:** Conceptualization, Methodology, Writing- Reviewing and Editing. **Carmen Prestemon:** Conceptualization, Methodology, Writing- Reviewing and Editing. **Cristina Lee:** Formal analysis, Writing- Reviewing and Editing. **Marissa Hall:** Conceptualization, Methodology, Writing- Reviewing and Editing, Funding acquisition.
